# Cortical Network Reorganization During Balance Control in Chronic Stroke: Premotor-Parietal Behavioral Uncoupling and Directional Prefrontal Compensation

**DOI:** 10.64898/2026.09.08.26362085

**Authors:** Komal K. Kukkar, Tina M. Pham, Humza A. Mirza, Sheng Li, Pranav J. Parikh

**Affiliations:** Center for Neuromotor and Biomechanics Research, Department of Health and Human Performance, University of Houston, Houston, Texas; Department of Physical Medicine and Rehabilitation, University of Texas Health Science Center – Houston, Houston, Texas

**Keywords:** Falls, network, EEG, dynamics, cortex

## Abstract

**Background:** Balance control is essential for independence in activities of daily living, and its impairment is a leading contributor to falls in stroke survivors. Stroke-induced brain reorganization alters cortical activation patterns; however, localized changes do not reveal the underlying network dynamics critical for the exchange of task-related information. How stroke affects balance-related cortical network dynamics remains unclear.

**Methods:** We studied cortical functional connectivity during a challenging balance task in fifteen chronic stroke survivors and fifteen age- and gender-matched healthy adults. Whole-head electroencephalography (EEG) was recorded while participants performed a balance sway-referenced task. Balance-related cortical sources were localized to the supplementary motor area/premotor area (SMA-PM), posterior cingulate cortex (PCC), and bilateral lateral prefrontal cortex (LPFC). Directed task-related functional connectivity was estimated across multiple frequency bands.

**Results:** Stroke participants demonstrated elevated gamma-band SMA-PM to PCC connectivity, ipsilesional LPFC to SMA-PM connectivity, and SMA-PM to contralesional LPFC connectivity compared to healthy participants. Stroke participants also showed poorer performance on both the clinical Berg Balance Scale (BBS) and the laboratory-based balance task. Stronger SMA-PM to contralesional LPFC connectivity, but not SMA-PM to PCC connectivity, was associated with better BBS scores in stroke survivors.

**Conclusions:** These findings suggest less reliable SMA-PM to PCC sensorimotor pathways and a compensatory recruitment of SMA-PM to contralesional LPFC pathway for the control of balance post-stroke. Cortical network reorganization after stroke leads to directional connectivity changes adaptive to functional needs, with implications for designing targeted interventions to reduce fall risk in stroke survivors.

## INTRODUCTION

Balance control requires the continuous integration of visual, vestibular, and somatosensory inputs to assess the body’s stability and generate anticipatory and/or corrective neuromuscular responses to prevent a fall [1–3]. In the healthy brain, this sensorimotor behavior is supported by functional connectivity within a distributed network, primarily linking posterior parietal areas and motor areas [4,5]. When the nervous system is compromised by aging or neurological injury, prefrontal cortical regions are recruited, and the reciprocal connection between prefrontal and motor areas may become important for monitoring ongoing balancing movements [6]. The premotor areas, such as the supplementary motor area, may serve as a hub that mediates interactions between sensorimotor and cognitive networks [7,8]. The coordination of information between cortical regions relies on frequency-specific neural oscillations [9–13]. Short-range synchrony in beta and gamma bands supports local processing [14,15], long-range synchrony in delta, theta, and alpha bands coordinates information flow between distant brain regions [12,16,17].

Stroke disrupts this coordinated frequency-specific cortical communication [18–20]. The structural disconnection of white matter pathways due to a stroke lesion directly or indirectly alters functional connectivity, leading to behavioral deficits [18,21–24]. Stroke-induced reorganization is reflected by reduced alpha and beta band connectivity in sensorimotor networks alongside increased gamma-band connectivity, representing a shift from long-range integrated to local segregated network organization [25]. The reorganization poststroke is observed in both hemispheres; it may be lateralized to the contralesional hemisphere initially, with a shift toward the ipsilesional hemisphere as functional recovery takes place [22,26,27]. Although stroke-related reorganization of cortico-cortical communication has been described for upper limb, how it affects the reorganization of cortico-cortical communication to support whole-body behaviors like balance remains poorly understood [28–30]. Functional connectivity patterns for lower limb control differ from those of the upper limbs. While upper limb control is heavily lateralized and discrete, postural stability is a bilateral task requiring distributed, whole-head cortical network coordination [31].

Neuroimaging studies have shown that stroke-induced cortical reorganization affects the activation of cortical brain areas during balance [5,32,33]. However, localized changes in the spectral power or metabolic activation patterns do not reveal the underlying network dynamics. A brain area can be highly active yet functionally segregated from the broader network [34]. Investigating functional connectivity and its changes due to stroke is required to determine how these sensorimotor and cognitive networks communicate to maintain balance stability, and how disruptions to these communications impair balance performance. Capturing these task-dependent frequency-specific connectivity requires source-space neuroimaging to understand the underlying cortical structures.

To address this gap, this study investigated source-localized cortical functional connectivity between regions activated during a challenging continuous balance task in chronic stroke survivors when compared with age-range-matched healthy adults. We recorded whole-head electroencephalography while participants performed a challenging sway-referenced balance task [2,35]. We hypothesized that, compared with healthy controls, stroke participants would demonstrate greater maladaptive gamma-related functional connectivity in the sensorimotor networks important for the balance task. We hypothesized that to compensate for the disrupted sensorimotor network, stroke participants would demonstrate an increased bilateral engagement of the prefrontal cortices. We expected that these connectivity patterns would be differentially associated with balance in stroke participants and healthy controls.

## METHODS

### Participants

Thirty adult participants provided written informed consent to participate in this study (**Table 1**). Fifteen participants had a history of stroke (60.8±9.69 years, mean ±SD; 37-70 years; Females=2) and fifteen participants served as age-range matched healthy controls (57.2 ±7.98 years, 35-70 years; Females=2). The inclusion criteria for participants with stroke included middle cerebral artery (MCA) stroke for the first time, at least 12 months post-stroke, ability to stand for 5 minutes independently without assistance, between 18 and 90 years of age, and a MoCA score ≥ 26 [36,37], and an absence of other neurological/ musculoskeletal impairments. Healthy adults (controls) had no history/symptoms of neurological/neuromuscular disorders affecting lower limbs with a MoCA score of 26 or greater. Healthy adults were dominant on the right foot. This study was approved by the Institutional Review Board at the University of Houston.

**Table 1.**
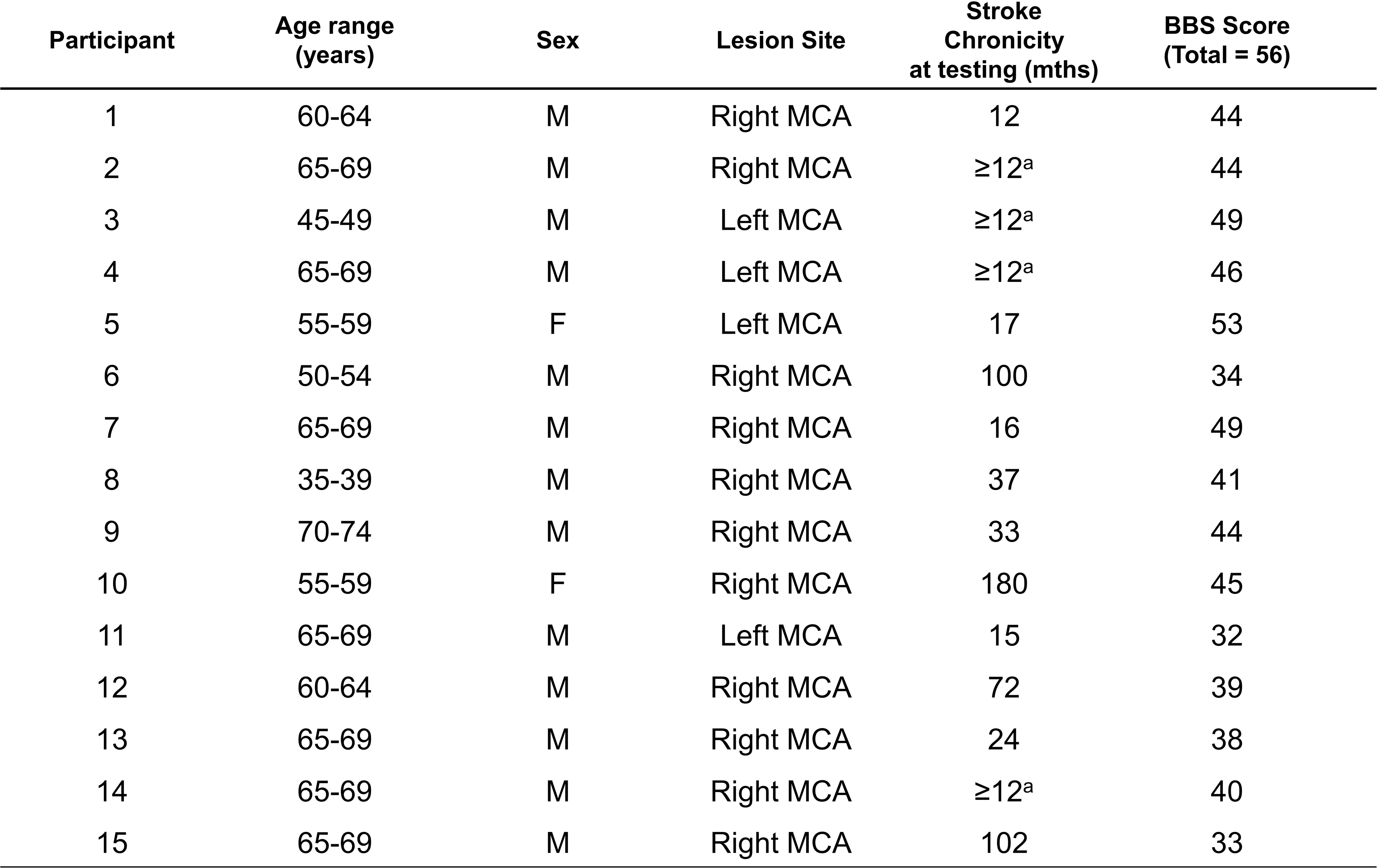
Patient demographics, characteristics, and clinical profile. ^a^exact data is not available; ^b^test was performed with an ankle-foot orthosis; F: female; M: male; MCA: middle cerebral artery; BBS: Berg Balance Scale.

| Participant | Age range<br>(years) | Sex | Lesion Site | Stroke<br>Chronicity<br>at testing (mths) | BBS Score<br>(Total = 56) |
| --- | --- | --- | --- | --- | --- |
| 1 | 60-64 | M | Right MCA | 12 | 44 |
| 2 | 65-69 | M | Right MCA | ≥12<sup>a</sup> | 44 |
| 3 | 45-49 | M | Left MCA | ≥12<sup>a</sup> | 49 |
| 4 | 65-69 | M | Left MCA | ≥12<sup>a</sup> | 46 |
| 5 | 55-59 | F | Left MCA | 17 | 53 |
| 6 | 50-54 | M | Right MCA | 100 | 34 |
| 7 | 65-69 | M | Right MCA | 16 | 49 |
| 8 | 35-39 | M | Right MCA | 37 | 41 |
| 9 | 70-74 | M | Right MCA | 33 | 44 |
| 10 | 55-59 | F | Right MCA | 180 | 45 |
| 11 | 65-69 | M | Left MCA | 15 | 32 |
| 12 | 60-64 | M | Right MCA | 72 | 39 |
| 13 | 65-69 | M | Right MCA | 24 | 38 |
| 14 | 65-69 | M | Right MCA | ≥12<sup>a</sup> | 40 |
| 15 | 65-69 | M | Right MCA | 102 | 33 |

### Instrumentation

#### Computerized dynamic posturography (CDP)

A commercially available CDP Force platform (Neurocom Balance Master, Natus Medical Incorporated, Pleasanton, CA) was used to assess dynamic balance stability (**Fig. 1A**). It is extensively used both in clinical [38] and research settings [39] for monitoring sensory and motor performance aspects of the balance control system. The platform is equipped with a motorized dual force plate system (45.72 cm x 45.72 cm), in which ground reaction forces (GRF) from under the subject’s feet are collected by normal and shear force transducers embedded within the force plate (support surface). The system can be used to adjust the orientation of the force plate with respect to the gravitational vertical by rotating it in the sagittal plane about an axis through the subject’s ankle joint in some proportion (a pre-selected gain between ‡2 and +2) to the postural sway of the subject. A negative sway gain means the movement of the plate will be in the opposite direction of the subject’s COP, and a positive sway gain means the movement of the plate will be in the same direction as the subject’s COP. The transducer data were collected at 100 Hz and processed by pre-installed software on a Windows-based desktop connected to the Neurocom Balance Master (Research module, Neurocom software version 8.0, Natus Medical Incorporated, Pleasanton, CA). The Neurocom system also generated an analog timing signal, which was used to synchronize the electroencephalography (EEG) system with GRF data.

**Figure 1.**
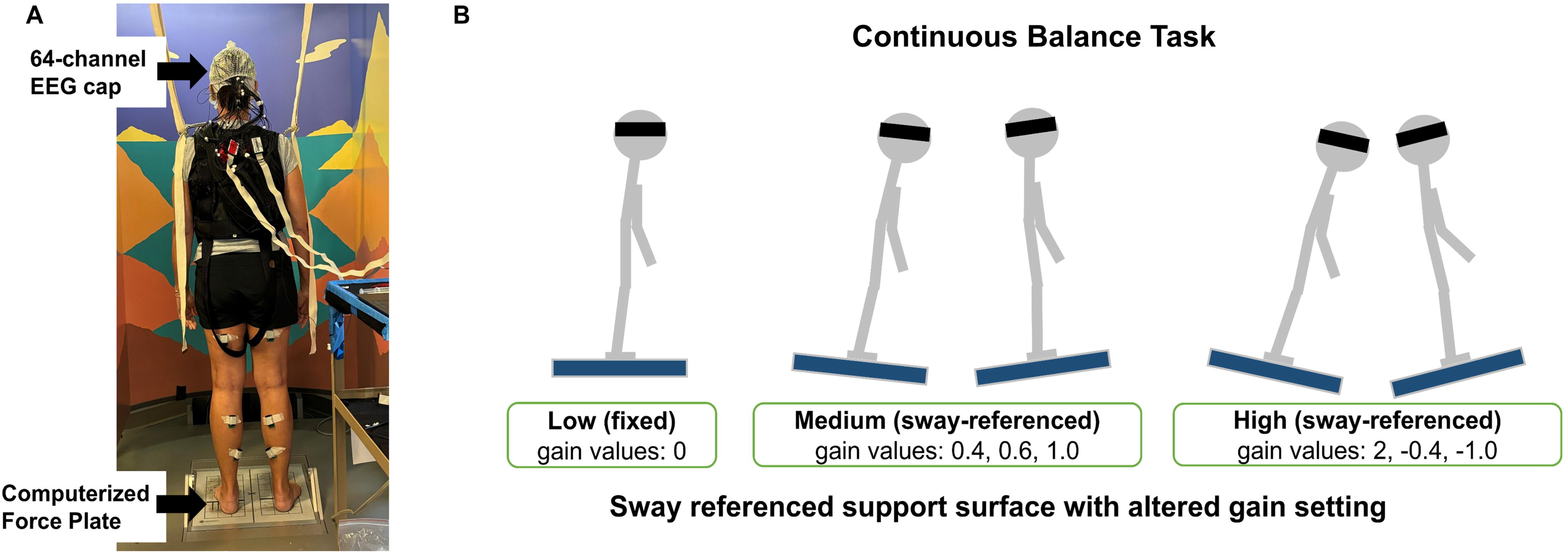
Experimental setup and continuous balance task. A. Experimental setup for the posture task. B. Continuous balance task. Schematic description of experimental conditions during the continuous balance task.

#### Berg Balance Scale (BBS)

It consists of a series of 14 predetermined tasks, and each task is scored, ranging from 0 to 4; “0” and “4” indicate the lowest and the highest functional levels, respectively. The score can take a value between 0 and 56, with higher scores suggestive of better balance ability [40,41].

#### Continuous Balance Task

Participants stood on the balance platform with eyes closed and were instructed to maintain their balance while EEG and GRF data were collected (**Fig. 1A**). The balance context was changed by varying the responsiveness of the support surface in proportion to the estimated center of mass sway (sway-referenced), which is intended to reduce the contribution of lower limb somatosensory receptors to the control of dynamic balance [42], and thereby manipulated the reliance on somatosensory inputs for the control of ongoing stance and modified the feedback relationship within the balance control loop. The effects of such manipulation on dynamic stability are related to the action of the participant. We achieved this by varying the gain of the support surface in the sagittal plane in different proportions to the estimated instantaneous COM sway angle. We used a range of gains (‡1.0, ‡0.4, 0, 0.4, 0.6, 1.0, 2.0) (**Fig. 1B**) in the balance task of three different levels of difficulty: *low*, *medium,* and *high*. The lowest gain value of 0 (fixed support surface with no sway referencing) may not be challenging to subjects and therefore was classified as *low* difficulty. Higher gain values of 0.4, 0.6, and 1.0 were classified as *medium* difficulty, as subjects were expected to exhibit greater balance instability. However, the gain of 2.0, or negative gains of ‡0.4 and ‡1.0, were classified as of *high* difficulty due to expectations that participants would have a further increase in balance instability, as done in our previous work [2,33,43]. The continuous balance task lasted for 180 s (20 s per gain), in the following order: 0, 0.4, 2, 0, 0.6, ‡0.4, 0, 1, ‡1. Participants were not informed when the gain changed. To ensure safety, participants always wore safety harnesses and a physical therapy belt with a spotter standing nearby to prevent any falls. Participants were allowed to practice at gain values of 0, 0.4, and 2 before exposure to the start of the data collection. Data from these gain values were not used for analysis.

#### Electroencephalography (EEG)

64 active channel EEG electrodes (Brain Products GmbH, Germany; 1000Hz) were used to record whole-scalp electroencephalography (EEG). The electrodes were placed as per the International 10-20 system for EEG electrode placement. Four of the electrodes were used to record electrooculography (EOG) signals. We constrained the movement of the EEG cables by placing an elastic mesh on the top of the EEG cap to minimize motion artifacts during the balance task on the Neurocom.[2]

#### Experimental Protocol

During this single-session study, we obtained participants’ clinical measures using the BBS test. Each participant then performed the continuous balance task while EEG was measured. The dynamic balance task consisted of maintaining an upright stance with eyes closed during nine sequential balance task conditions (gains), each lasting for 20 sec.

### Data Analyses

#### Behavior

BBS was scored out of 56 total points. The ground reaction force data were used to create a center of pressure (COP) time series for the balance task [44]. Linear and non-linear measures of postural performance were computed: root mean square COP (RMS COP) [45], path length (PL) [46] from COP [47], RMS COP velocity (RMS COPv) [48], and the sample entropy (SE) of COP [49]. The number of falls during the balance task was recorded during testing. A fall was noted when a participant lost balance and required either self-induced stepping and/or support from the overhead harness to prevent them from falling to the floor.

#### EEG

The pre-processing pipeline used in this study was similar to that described in our earlier studies [35,50]. The EEG data were downsampled to 250 Hz. An artifact-removal H-infinity filter was applied to remove ocular artifacts and signal drifts [51]. The eye-blink and eye-motion artifacts were filtered using data recorded from EOG channels around the eyes as a reference signal [52]. A Zapline function in EEGLab was used to remove the line noise [53]. A standardized early-stage EEG processing pipeline (PREP) with default parameters was used to remove artifactual EEG channels and apply a robust common referencing method to increase the signal-to-noise ratio [54]. The PREP pipeline also replaces artifactual channels with surrogate data that is interpolated from neighboring sensors to minimize bias when performing common average referencing. The signals were then band-pass filtered at 0.1-100 Hz using a 4th-order Butterworth filter to remove slow drifting noise. Artifact Subspace Reconstruction (ASR) was applied next to detect and denoise any artifactual sections in the EEG data [55–57]. Adaptive mixture ICA (AMICA) was used to compute the maximally independent components (ICs) from the data [35,50]. The AMICA is reported to be the best ICA method available to date, and it provides more dipolar and minimized mutual information ICs [58,59]. Using the constructed boundary elemental model (BEM) explained in the next paragraph and the digitized channel locations, the dipole fitting method: *DIPFIT* in EEGLAB [60,61] was used to calculate the equivalent current dipole sources that explain at least 85% [62] of topographic variance obtained from ICA results. The digitized channel locations were first warped onto the constructed head model before calculating the dipole locations. We removed dipoles located outside the individual BEM model, as well as those with artifactual components such as muscle-related power spectral density characteristics and motion artifact-related high-frequency noise. The boundary element model (BEM) was created after computing surface meshes and constructing the head model using a standard MRI template. Each IC scalp projection, its equivalent dipole’s location, and its power spectra were then visually inspected, and ICs that related to non-brain artifacts (e.g., motion, muscle artifact) were removed.

After artifact removal and dipole fitting, we had 298 ICs remaining for the stroke group and 238 ICs remaining for the healthy control group. To obtain common sources across both groups, we combined the ICs from all stroke and healthy participants, normalized to the MNI (Montreal Neurological Institute, Quebec, Canada). Independent components were clustered into six groups using k-means clustering based on 3D dipole locations, only to avoid circular interference, which is known to cause an increased false positive rate [63,64]. The number of clusters (k = 6) was chosen to ensure a comprehensive representation of each subject’s dipoles while minimizing the risk of data overfitting [65,66]. Using the same variance criteria as above, keeping only those dipoles that accounted for at least 85% of variance and were within 3 SD of the cluster centroid, brought the number of ICs from 536 to 328, which were used for clustering. Clustering was performed using our own customized MATLAB application named EEG_Clustering (https://github.com/kkukka21/EEG_Clustering), converted to an EEGLAB plugin extension for group-level analysis and visualization of spatiotemporal properties of ICs. The tool serves as an alternative to the EEGLAB STUDY framework by enabling IC clustering based on estimated dipole coordinates. The clusters included independent components from at least 60% of participants, suggesting the clusters were representative of most of the participants in each group. The Brodmann Area (BA) for each cluster was determined using the Talairach Daemon software as an area within 1 mm of the cluster centroid [67,68]. Common clusters across the two groups were identified as sources of EEG activations demonstrating similar Brodmann areas.

Following the clustering analysis, we estimated multivariate Granger Causality (GC) between cluster dipoles using the multivariate GC Toolbox, as done in our previous work [5]. MVGC Toolbox operationalizes G-causality through vector autoregressive (VAR) modeling and captures dependencies among multivariate time series. This modeling approach is well-suited for analyzing the dynamic interactions of neural signals, such as EEG data with high temporal resolution and stochastic nature. Granger causality assesses causality based on the principle that a cause precedes and predicts its effect, thereby estimating directed interactions [69,70]. Consider two jointly distributed vector-valued stochastic processes X and Y. If the past of Y conveys information about the future of X above and beyond all information contained in the past of X, then we say that Y G-causes X. The VAR model underlying GC estimation is expressed as [71,72]

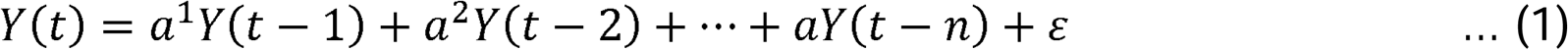

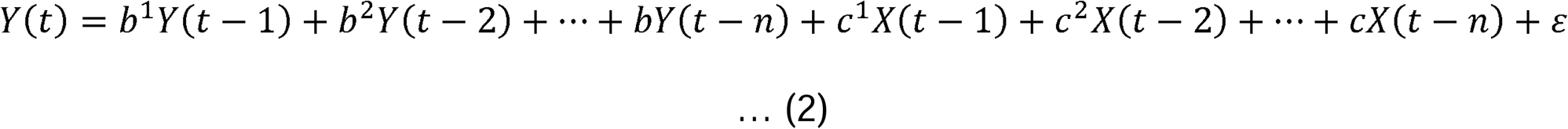

where Y(t) is the time series being predicted, X(t) is the time series used for prediction, a□ and b□ are the autoregressive coefficients, c□ are the coefficients from the X(t) time series, and ε⍰ denotes the residual error (unexplained variance). Equation (1) represents the restricted model where Y is predicted only by its own past, while equation (2) represents the unrestricted model that includes the past of X as an additional predictor. The Granger causality from X to Y is then quantified as the logarithmic ratio of the prediction error variances[73]:

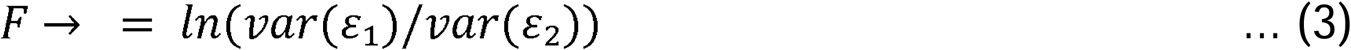

where var(ε₁) is the variance of residuals from the restricted model (Equation 1) and var(ε₂) is the variance of residuals from the unrestricted model (Equation 2). If F□→□ > 0, then X is said to Granger-cause Y, indicating that including the past of X significantly improves the prediction of Y’s future. However, if there is no direct causal influence of Y on X, but both X and Y depend on another variable Z, this can result in spurious Y-to-X causality. To address this, we employed pairwise-conditional G-causality to estimate the connectivity between two clusters while conditioning on all other identified clusters. This approach eliminates spurious causalities by “conditioning out” common dependencies when they are available in the data. The connectivity analysis was conducted for each subject using dipoles that contributed to the clusters. The dipole time series were first epoched into individual balance task conditions (low, medium, and high). Granger Causality analysis strictly requires the time series data to be stationary. The Augmented Dickey-Fuller test was conducted on each dipole time series for each participant to determine whether the dataset is stationary. For each subject, dipole-to-dipole GC coefficients were estimated for each condition in the frequency domain across the following bands: delta (0.5 to <4 Hz), theta (4 to <8 Hz), alpha (8 to <12 Hz), beta (12 to <30 Hz), and gamma (30 to <50 Hz). The optimal VAR model order was determined for each participant via the Akaike Information Criterion (AIC) to ensure the framework accurately captured spectral dynamics. Higher EEG functional connectivity coefficients indicate stronger connectivity [69,70]. We excluded subject data from clusters for which individual subjects did not contribute any independent component. Additionally, we removed trials during which subjects required assistance.

#### Statistical Analyses

For BBS scores, these assumptions were found to be violated (p<0.05), and thus, a non-parametric approach, the Mann-Whitney U test, was used for group comparison. For data collected during the continuous balance task, the first three practice conditions and the trials were participants fell/required assistance were not included in the analysis. The last three trials had more participants who received assistance in the form of a step or from a spotter, leading to the deletion of more than 30% of data. Therefore, these trials were also excluded. For analysis, we use trials with 0 (low), 0.6 (medium), and ‡0.4 (high) gain settings (20 sec each). We performed normality (Shapiro-Wilk) and equal variance Levene’s test to check for violation of assumptions for the parametric statistical approach.

For each COP measure, we used maximum likelihood linear mixed models with between-subject factor as Group (Stroke, Healthy) and within-subject factors such as Task Condition (Low, Medium, High). The source-level EEG functional connectivity coefficients between all possible pairwise combinations of the clusters of interest (source-level) were used.

For each EEG cortico-cortica<u>l</u> connectivity pair (SMA-PM ⇄ PCC; Ipsi/Dom LPFC ⇄ SMA-PM; Contra/Non LPFC ⇄ SMA-PM; PCC ⇄ Ipsi/Dom LPFC; PCC ⇄ Contra/Non LPFC), we used maximum likelihood linear mixed models with a between-subject factor as Group (stroke, healthy) and within-subject factors such as Connectivity Direction (from/to region), Task Condition (low, medium, and high), and Frequency Bands (delta, theta, alpha, beta, and gamma). Because stroke participants may utilize distinct functionally connected networks for balance when compared with healthy controls, we performed separate correlation analyses within each group. Spearman’s rank correlation analysis was used to examine the relationship between the functional connectivity coefficients and clinical Berg Balance scores (the sum of ordinal items). Sidak-Bonferroni correction for multiple comparisons was used to control statistical errors. The significance level α was set at 0.05 (IBM SPSS v29.0).

## RESULTS

We found no age-related difference between the two groups (t_28_= 1.11; p=0.28) and no group differences in the number of trials eliminated (Stroke:9%; Healthy:7%; χ^2^: df=1, 2.36; p>0.05). Only one participant in the stroke group had both medium and high balance task condition trials excluded.

### Behavior

As expected, stroke participants (42.07 ±6.16; mean ±SD), when compared to healthy controls (54.73 ±1.80), showed lower (i.e., poor) scores on the Berg Balance Scale (**Fig. 2A**; Mann-Whitney U-test; *U*=2.5, *Z* =– 4.606; *p*<0.001).

**Figure 2.**
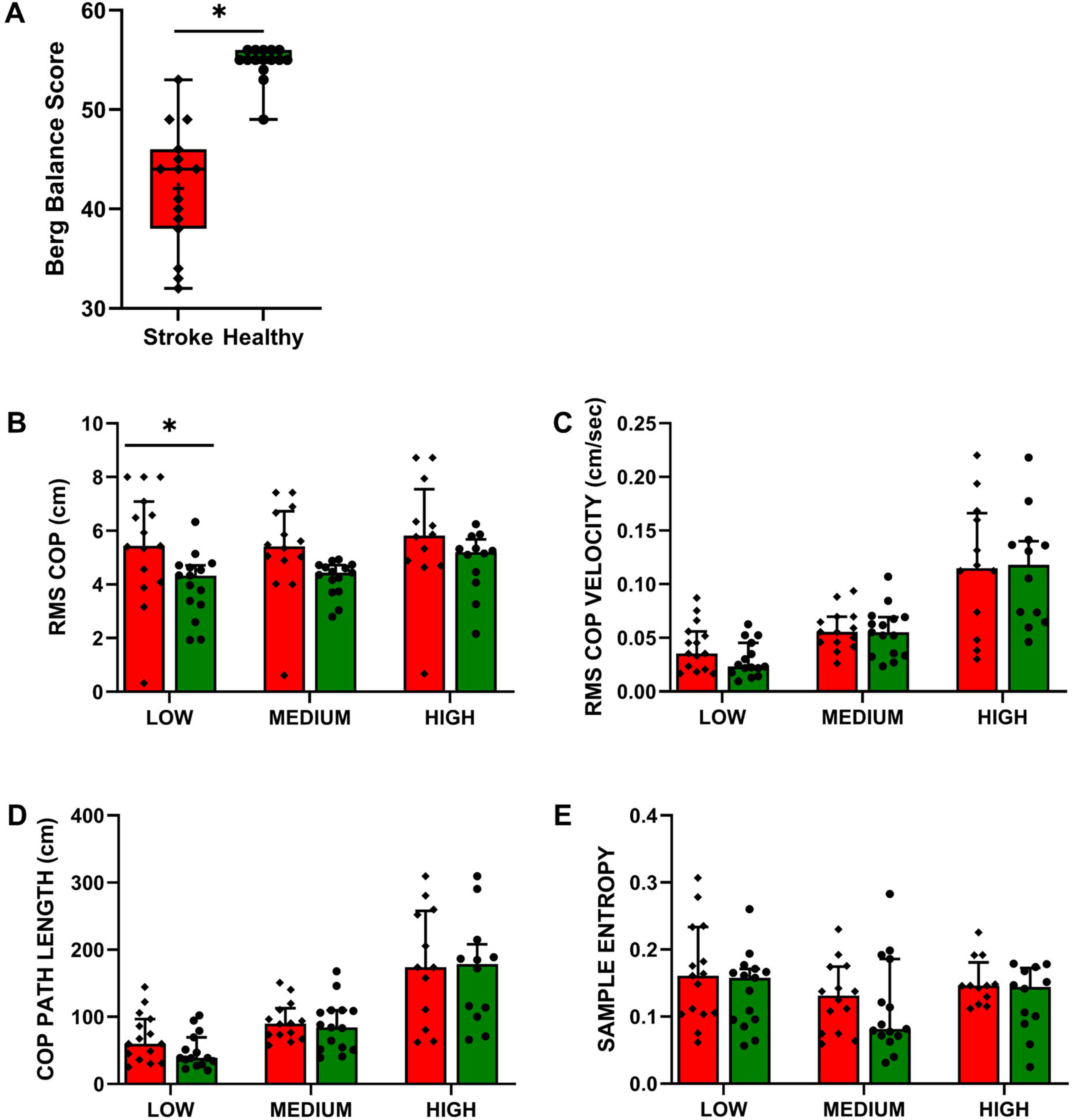
Behavioral variables. A. Berg Balance Score (BBS) shown as median and quartiles. B. RMS COP in the antero-posterior direction measured during the continuous balance task, C. RMS COPv in the antero-posterior direction, D. COP Path length (PL) E. Sample Entropy of COP. COP data are mean (± SEM) of all subjects. Asterisks indicate significant differences between groups.

We found higher RMS COP in stroke than in healthy controls (**Fig. 2B**; Omnibus multi-factor linear mixed model: Group: F_1,9.3_=5.902, p=0.037). We also found a significant increase in RMS COP with increasing difficulty of the balance task conditions (Condition: F_2,26.03_=8.97, p=0.001). However, this increase was similar across both groups (no Group × Condition interaction: F_2,26.03_=0.619, p=0.546). Post hoc pairwise comparisons were non-significant (all p>adjusted α).

RMS COPv increased with an increase in the difficulty of the balance task conditions; however, this increase was similar across groups (**Fig. 2C**; Omnibus multi-factor linear mixed model: Condition: F_2,68.79_=65.99, p<0.001; no Group x Condition interaction: F_2,68.79=_0.334, p=0.717, and Group: F_1,65.34_=0.334, p=0.565). Post hoc pairwise comparisons were non-significant (all p>adjusted α).

Similarly, COP PL increased with an increase in the balance task difficulty conditions; however, this increase was similar across groups (**Fig. 2D**; Omnibus multi-factor linear mixed model: no Group × Condition interaction: F_2,69.67_=0.305, p=0.738; Condition: F_2,69.67_=65.69, p<0.001, and Group: F_1,80.91_=0.616, p=0.435). Post hoc pairwise comparisons were non-significant (all p>adjusted α).

The sample entropy, a non-linear COP measure, also modulated significantly across the balance task difficulty conditions; however, there was no group difference (**Fig. 2E**; Omnibus multi-factor linear mixed model: Condition: F_2,55.15_=3.631, p=0.033; no significant Group × Condition interaction: F_2,55.15 =_ 0.367, p=0.694, and Group: F_1,44.81_=0.116, p=0.735). Post hoc pairwise comparisons were non-significant (all p>adjusted α).

### EEG Connectivity

The number of participants contributing to each cluster ranged from 18 (60%) to 28 (93%), indicating consistent identification of these sources across the study samples (**Fig. 3A and 3B**). Cluster 1 was localized to the supplementary motor area-premotor area (SMA-PM), Cluster 2 was positioned in the posterior cingulate cortex (PCC), Cluster 3 was localized to the right lateral prefrontal cortex (right LPFC), and Cluster 4 was positioned in the left lateral prefrontal cortex (left LPFC). In addition, we found clusters localized within the left and right occipital gyri. Because this study focused on interactions between sensorimotor and prefrontal networks, we focused our analysis and discussion on SMA-PM, PCC, and bilateral LPFC.

**Figure 3:**
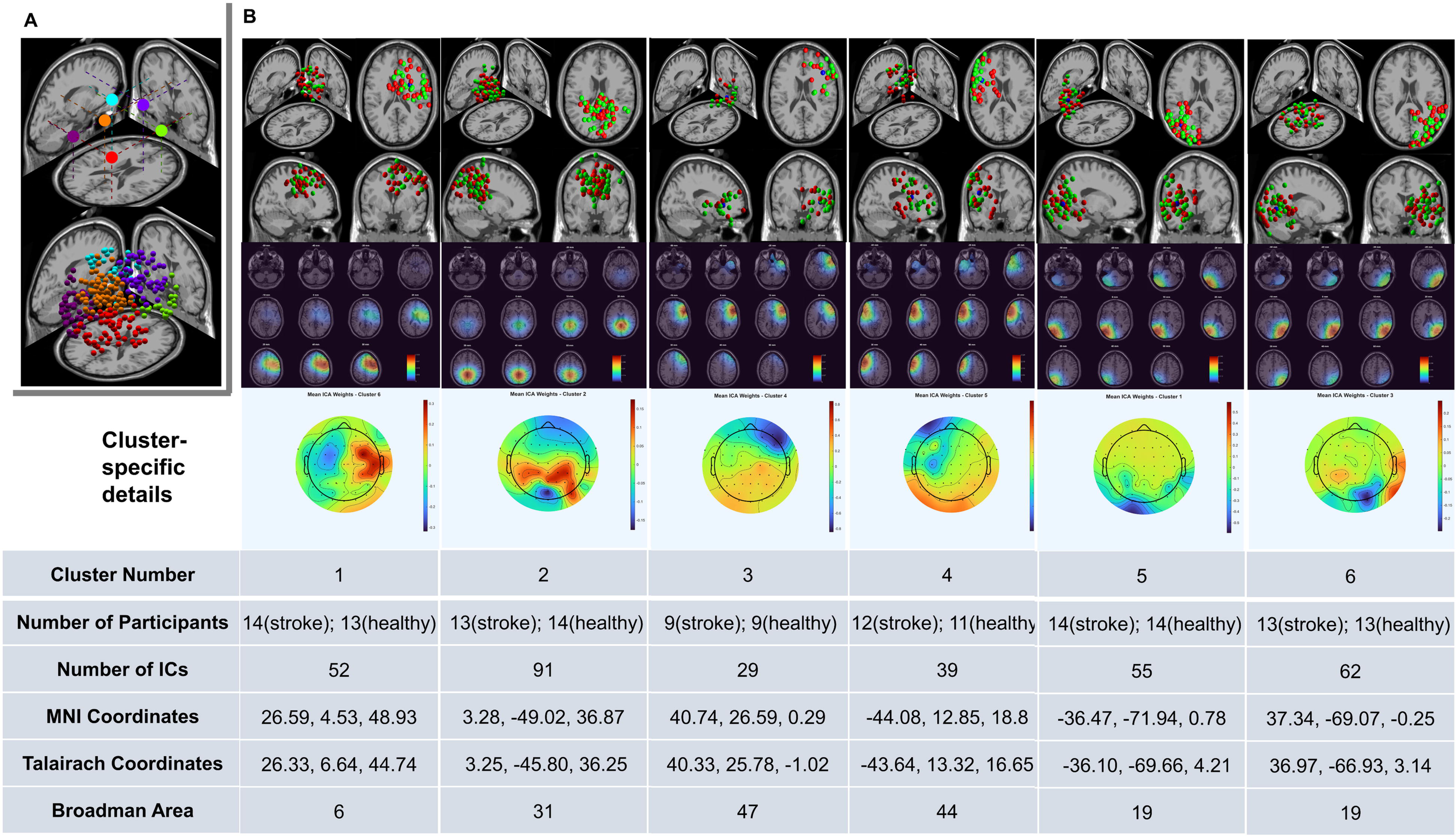
Source-localized cortical regions of activation during the continuous balance task. A. (top) Centroids of all clusters identified across participants, displayed on an oblique view of a brain slice. Each colored sphere represents the centroid location of one of the six identified clusters. (bottom) Three-dimensional dipole locations of all ICs are shown on the same oblique brain slice view. Dipoles are color-coded to match their corresponding cluster centroids in panel A, illustrating the anatomical spread of sources within each cluster across participants. B. Brain clusters. (top) Four orthogonal views (oblique, axial, sagittal, and frontal) of each cluster showing anatomical localization with group-specific color coding: red = stroke participants, green = healthy participants, blue = cluster centroid. (middle) Cluster density plots displaying axial slices from ‡50 mm to +50 mm (inferior to superior), with color intensity indicating the concentration of ICs at each depth level. (bottom) Scalp topography maps for each cluster showing the scalp projection patterns of the corresponding sources. The table summarizes cluster properties, including participant composition (stroke versus healthy), number of ICs, MNI coordinates (x, y, z in mm), Talairach coordinates (x, y, z in mm), and corresponding Brodmann area with anatomical labels. Brodmann areas are the regions found within a ±5 mm search range of the cluster centroids.

The Augmented Dickey-Fuller test rejected the null hypothesis of a unit root (all ADF test statistics <-51.2; all p<0.01), demonstrating that all dipole time series for all participants were stationary. To account for lesion lateralization, EEG clusters were aligned such that the hemisphere ipsilateral to the stroke lesion was designated as ipsilesional. For the source-localized functional connectivity analysis, we focused on functional connectivity between four regions: PCC, bilateral LPFC, and SMA-PM. For comparison, ipsilesional LPFC (Ipsi) in stroke participants was paired with LPFC in the dominant (Dom; the ipsilesional equivalent) hemisphere for healthy controls. Contralesional (Contra) LPFC was paired with LPFC in the non-dominant (Non; the contralesional equivalent) hemisphere for healthy controls. This resulted in five bidirectional pairwise functional connectivity measures: PCC ⇄ SMA-PM, Ipsi/Dom LPFC ⇄ SMA-PM, Contra/Non LPFC ⇄ SMA-PM, PCC ⇄ Ipsi/Dom LPFC, and PCC ⇄ Contra/Non LPFC.

### SMA-PM ⇄ PCC

Stroke participants demonstrated distinct frequency-dependent differences in SMA-PM ⇄ PCC EEG functional connectivity when compared with healthy controls (Omnibus multi-factor linear mixed model: significant Group × Band interaction: F_4,704.857_=2.764, p=0.027). The model also revealed a significant main effect of Direction (F_1,704.488_=19.343, p<0.001) and Band (F_4,704.857_=14.071, p<0.001). No other main effects or interaction effects reached statistical significance (all F<1.1; all p>0.3). Post hoc pairwise comparisons with Sidak-Bonferroni correction showed significantly greater EEG functional connectivity coefficients in the gamma frequency band from SMA-PM → PCC in stroke versus healthy participants (p=0.026; **Fig. 4A-B**). All other pairwise comparisons did not reach statistical significance (all p>adjusted α=0.025; **Fig. 4C-D**).

**Figure 4.**
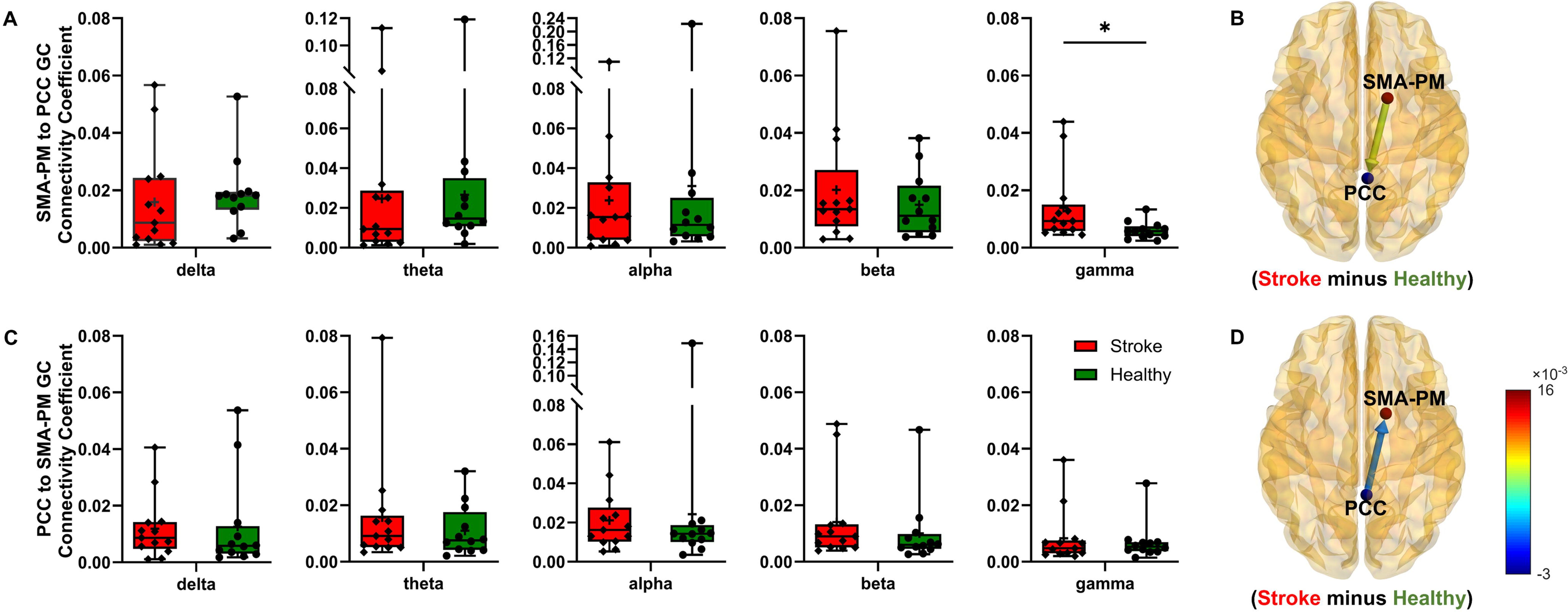
EEG functional connectivity between SMA-PM and PCC. A. and B. Box and whisker plot for each frequency band and brain visualization (stroke minus healthy) for the gamma frequency band, respectively, for the SMA-PM to PCC direction. Asterisk denotes statistical significance (*p < 0.05). C. and D. Box and whisker plot for each frequency band and brain visualization (stroke minus healthy) for the gamma frequency band, respectively, for the PCC to SMA-PM direction. Red boxes represent stroke participants; green boxes represent healthy controls. The color scale in panels B and D represents functional connectivity strength differences, with warmer colors indicating higher connectivity in stroke participants.

### Ipsi/Dom LPFC ⇄ SMA-PM

Stroke participants demonstrated significant differences in EEG functional connectivity coefficients across directional pathways when compared to healthy controls (Omnibus multi-factor linear mixed model: significant Group × Direction interaction: F_1,119.248_=10.685, p=0.001; **Fig. 5A-D**). The model also revealed main effects of Condition (F_2,117.325_=6.038, p=0.003) and Band (F_4,119.415_=11.421, p<0.001). No other main effects or interaction effects reached statistical significance (all F<3.1; all p> 0.09).

**Figure 5.**
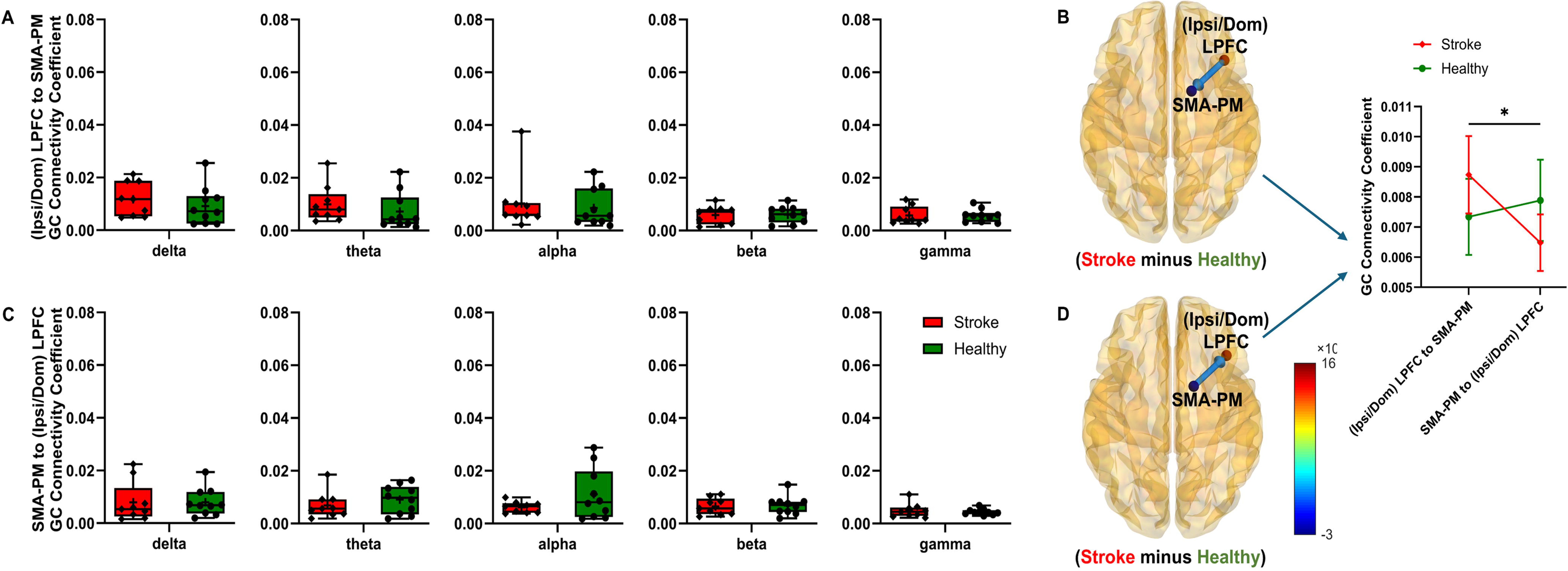
EEG functional connectivity between SMA-PM and Ipsilesional (Ipsi) LPFC in stroke participants and Dominant (Dom) LPFC in healthy participants. A. and B. Box and whisker plot for each frequency band and brain visualization (stroke minus healthy) averaged across frequency bands, respectively, for the Ipsi/Dom to SMA-PM direction; inset line plot showing significant Group × Direction interaction. Asterisk denotes statistical significance (*p < 0.05). C. and D. Box and whisker plot for each frequency band and brain visualization (stroke minus healthy) averaged across frequency bands, respectively, for the SMA-PM to Ipsi/Dom LPFC direction.

The absence of three-way or four-way interactions involving Frequency Band or Condition indicates that the observed directional network reorganization in stroke participants represents a generalized phenomenon across frequency bands that remains stable across varying balance task conditions. Post-hoc simple effects to tease out the significant Group × Direction interaction showed that Ipsi LPFC → SMA-PM EEG functional connectivity coefficients were significantly greater than those for SMA-PM → Ipsi LPFC in stroke participants (p<0.001; adjusted α=0.025). In contrast, in healthy controls, there was no difference between Dom (the ipsilesional equivalent) LPFC → SMA-PM and SMA-PM → Dom LPFC connectivity coefficients (p=0.22). No other post-hoc comparisons were significant (all p>0.05; **Fig. 5A-D**).

### Contra/Non LPFC ⇄ SMA-PM

Stroke participants demonstrated significant group differences in EEG functional connectivity coefficients across directional pathways compared with healthy controls (Omnibus multi-factor linear mixed model: significant Group × Direction interaction: F_1,627.600_=13.921, p<0.001; **Fig. 6A-D**). The model also revealed main effects of Condition (F_2,613.127_=4.988, p=0.007) and Band (F_4,627.558_=7.331, p<0.001), and a significant Direction × Condition interaction (F_2,627.636_=4.519, p=0.011). No other main effects or interaction effects reached statistical significance (all F<1.92; all p>0.1). The absence of three-way or four-way interactions involving Frequency Band or Condition indicates that the observed directional network reorganization in stroke participants represents a generalized phenomenon across frequency bands that remains stable across varying balance task conditions. Post-hoc simple effects testing to tease out the significant Group × Direction interaction showed that SMA-PM → Contra LPFC EEG functional connectivity coefficients were significantly greater than Contra LPFC → SMA-PM in stroke participants (p=0.01; adjusted α=0.025). In contrast, healthy controls showed significantly greater EEG functional connectivity coefficients from Non (the contralesional equivalent) LPFC → SMA-PM when compared with SMA-PM → Non LPFC (p=0.006). No other post-hoc comparisons were significant (all p>0.05; **Fig. 6A-D**).

**Figure 6.**
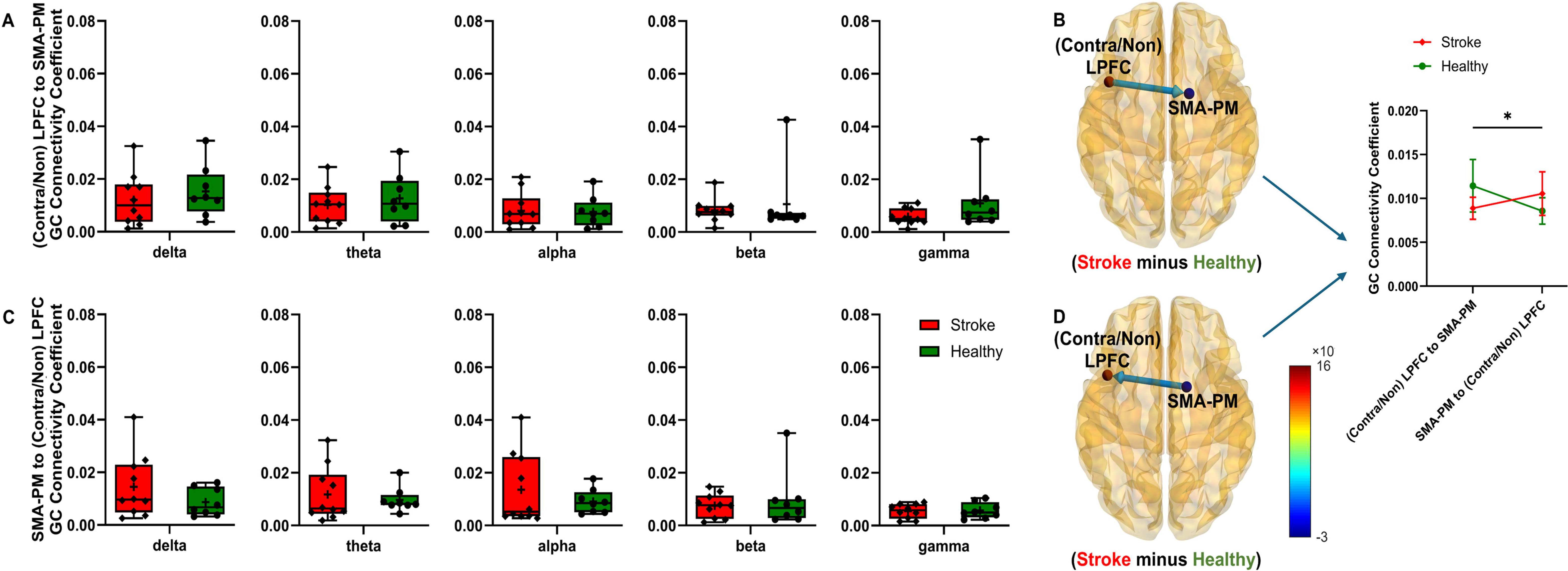
EEG functional connectivity between SMA-PM and Contralesional (contra) LPFC in stroke participants and Non-dominant (Non) LPFC in healthy participants. A. and B. Box and whisker plot for each frequency band and brain visualization (stroke minus healthy) averaged across frequency bands, respectively, for the Contra/Non to SMA-PM direction; inset line plot showing significant Group × Direction interaction. Asterisk denotes statistical significance (*p < 0.05). C and D. Box and whisker plot for each frequency band and brain visualization (stroke minus healthy) averaged across frequency bands, respectively, for the SMA-PM to Contra/Non LPFC direction.

### PCC ⇄ Ipsi/Dom LPFC

Stroke participants demonstrated significant group differences in EEG functional connectivity coefficients across frequency bands when compared to healthy controls (Omnibus multi-factor linear mixed model: significant Group × Band interaction: F_4,947.672_=3.762, p=0.005; Condition: F_2,943.524_=4.452, p=0.012; Band: F_4,947.672_=13.870, p<0.001; **Fig. 7A-D**). There was no Group × Direction interaction (F_1, 481.869_=0.01, p=0.92). No other main effects or interaction effects reached statistical significance (all F<2.1; all p>0.09). While the significant omnibus Group × Band interaction is suggestive of a generalized frequency-dependent change between groups, follow-up pairwise comparisons with Sidak-Bonferroni correction revealed that no individual frequency band differences reached the adjusted significance threshold (all p>0.05).

**Figure 7.**
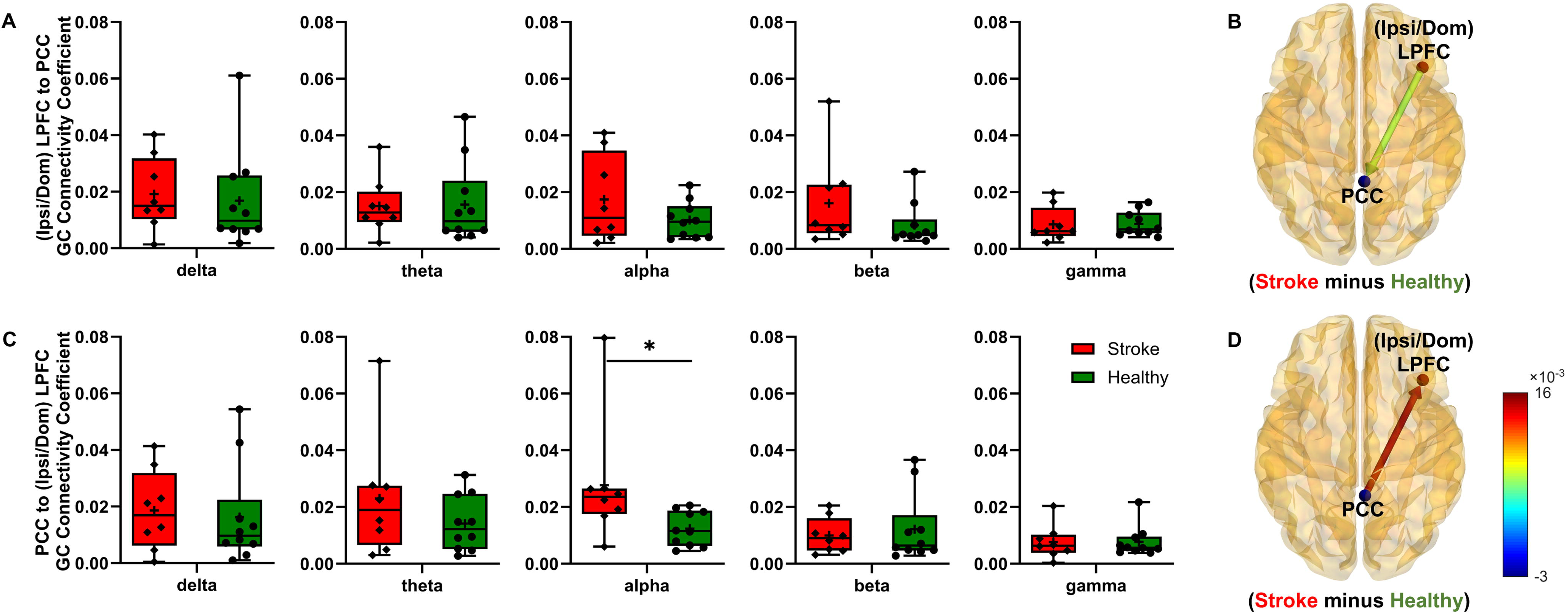
EEG functional connectivity between PCC and Ipsilesional (Ipsi) LPFC in stroke participants and Dominant (Dom) LPFC in healthy participants. A. and B. Box and whisker plot for each frequency band and brain visualization (stroke minus healthy) averaged across frequency bands, respectively, for the Ipsi/Dom LPFC to PPC direction. C. and D. Box and whisker plot for each frequency band and brain visualization (stroke minus healthy) averaged across frequency bands, respectively, for the PCC to Ipsi/Dom LPFC direction.

### PCC ⇄ Contra/Non LPFC

Stroke participants demonstrated significant group differences in EEG functional connectivity coefficients across directional pathways when compared to healthy controls (Omnibus multi-factor linear mixed model: significant Group × Direction interaction: F_1,195.551_=5.000, p=0.026; main effect of Band: F_4,195.654_=15.306, p<0.001; **Fig. 8A-D**). No other main effects or interaction effects reached statistical significance (all F<2.9; all p>0.1). While the significant omnibus Group × Direction interaction suggests a generalized directional shift in network topology between groups, follow-up pairwise comparisons revealed that no individual directional pathway differences reached the adjusted significance threshold (all p>0.05).

**Figure 8.**
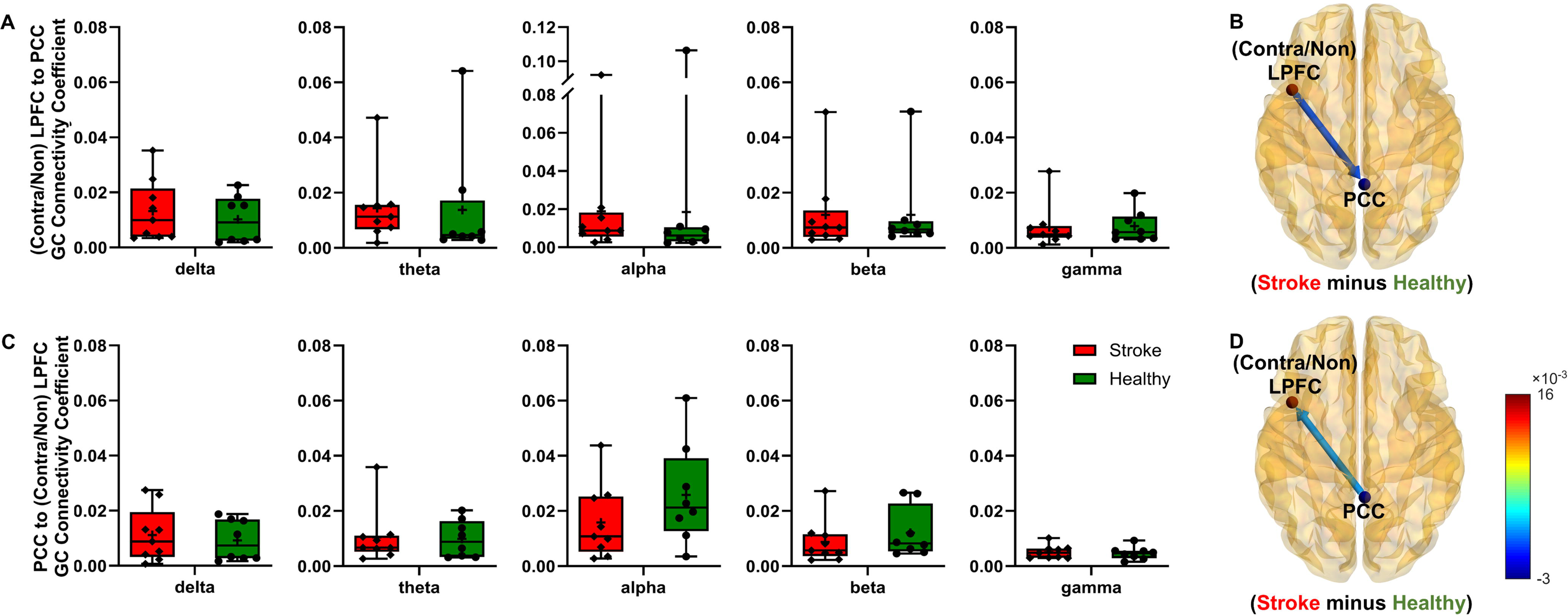
EEG functional connectivity between PCC and Contralesional (contra) LPFC in stroke participants and Non-dominant (Non) LPFC in healthy participants. A and B. Box and whisker plot for each frequency band and brain visualization (stroke minus healthy) averaged across frequency bands, respectively, for the Contra/Non LPFC to PPC direction. C and D. Box and whisker plot for each frequency band and brain visualization (stroke minus healthy) averaged across frequency bands, respectively, for the PCC to Contra/Non LPFC direction.

### Association between EEG functional connectivity and BBS in Stroke

Stronger SMA-PM → PCC functional connectivity coefficients in the gamma band did not correlate with BBS score in stroke participants (Spearman’s ρ=-0.02, p=0.96; **Fig. 9A**).

**Figure 9.**
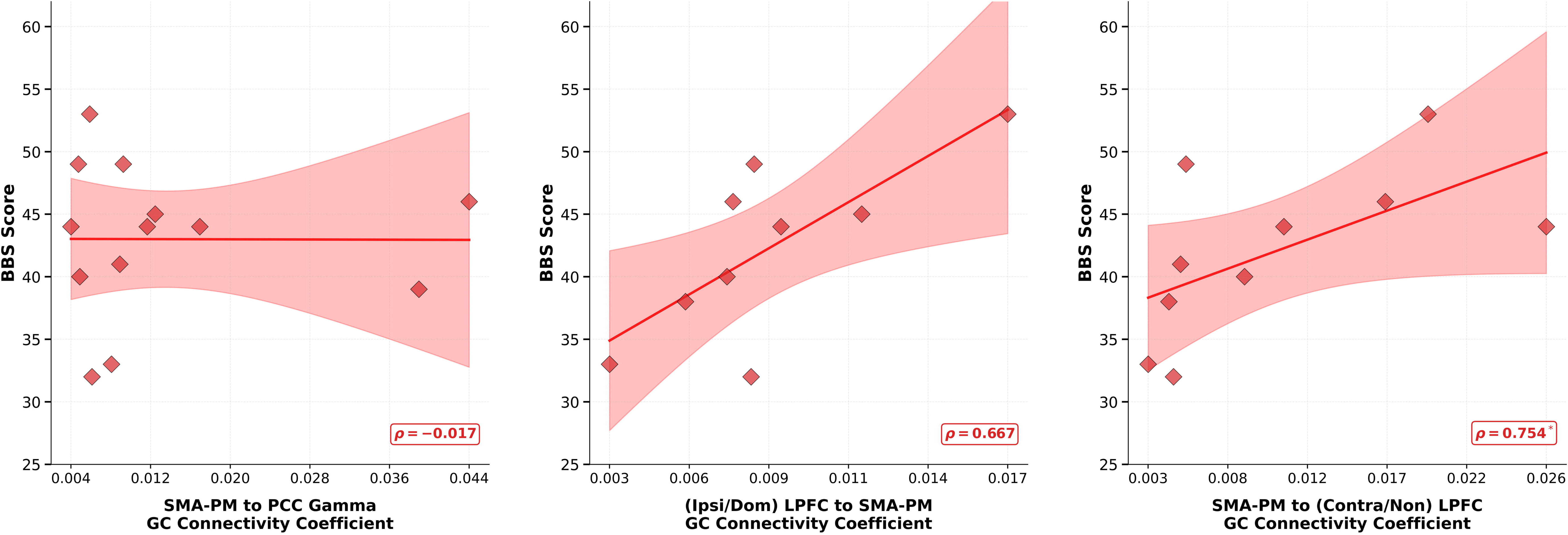
Association between EEG functional connectivity coefficients and Berg Balance score in stroke participants. A. No significant association was found between SMA-PM to PCC functional connectivity (anterior to posterior) in the gamma band (pooled across all conditions) and BBS score in stroke participants (p = 0.96). B. A correlation between higher Ipsi/Dom LPFC to SMA-PM functional connectivity (pooled across all bands and conditions) and better BBS score in stroke participants did not survive the adjusted significance threshold (p = 0.05 uncorrected; adjusted α = 0.025). C. Higher SMA-PM to Contra/Non LPFC functional connectivity (pooled across all bands and conditions) was associated with better BBS score in stroke participants. Red data points and corresponding line of fit represent stroke participants. Shaded bands represent 95% confidence intervals. Asterisk denotes statistical significance (*p < 0.05).

Given a significant Group × Directions interaction, we pooled data across all frequency bands and conditions to assess the correlation between the connectivity of SMA-PM with bilateral PFCs and BBS score. In stroke participants, stronger Ipsi PFC → SMA-PM functional connectivity was associated with higher (i.e., better) BBS score, but it failed to reach the adjusted significance threshold of 0.025 (Spearman’s ρ=0.68, p=0.05; **Fig. 9B**). Stronger functional connectivity from SMA-PM → contralesional LPFC was significantly correlated with better BBS score in stroke participants. (Spearman’s ρ=0.754, p=0.015; adjusted α=0.025; **Fig. 9C**).

## DISCUSSION

This study investigated source-localized directional functional connectivity during a challenging balance task in chronic stroke survivors and healthy adults. Across groups, SMA-PM, PCC, and bilateral LPFC were activated during performance of the balance task. The novel findings from this study are that stroke participants demonstrated elevated gamma-band SMA-PM → PCC connectivity, elevated ipsilesional LPFC → SMA-PM connectivity, and elevated SMA-PM → contralesional LPFC connectivity. The behavioral relevance of these networks differed in stroke participants, with no significant SMA-PM → PCC connectivity association with balance found, whereas SMA-PM → contralesional LPFC connectivity was associated with better balance ability. These findings suggest that distinct directional executive-sensorimotor pathways differentially support balance post-stroke.

### Elevated Gamma band SMA-PM → PCC Connectivity is Behaviorally Uncoupled After Stroke

PCC in the posteromedial cortex, which includes the parietal regions, is an important hub for multiple brain networks, including the default mode network, frontoparietal control network, salience network, and sensorimotor network [74]. Studies in nonhuman primates [75] and humans [76] suggest that PCC acts as a detector of unexpected events, triggering reactive mechanisms to adapt behavior to changing conditions. The SMA-PM region utilizes the sensory information received from PCC via reciprocal corticocortical fibers [77] for motor planning, action selection, and influences voluntary movements through projections to the primary motor cortex [78,79]. SMA-premotor and parietal regions of the brain are presumed to play a role in filtering out the sensory information arising from our own actions so that the system is attuned to external events for motor control [80–82]. fMRI and TMS studies suggest that SMA-PM areas generate a copy of the motor command, a feedforward signal, which is then used to estimate the sensory consequence of a planned movement [80,81]. Parietal regions may give rise to sensory prediction and compare it with the actual sensory information to suppress self-generated sensory events and highlight any externally generated sensory events, for example, unexpected platform movements due to the gain change. Any mismatch would then require sending the information back to SMA-PM for correction of the motor plan. In our study, the flow of information from SMA-PM → PCC connectivity may reflect transmission of a copy of the balance-related motor command, which PCC uses to maintain spatial orientation and keep the participant from falling during the continuous balance task [75,76,83,84]. This feedforward communication is thought to be mediated by synchronization in the gamma frequency range [85,86]. While stroke participants showed stronger SMA-PM → PCC connectivity in the gamma frequency band when compared with healthy adults, this elevated connectivity did not reflect increased recruitment for sensorimotor compensation as has been observed in earlier stroke work [87–89]. Instead, we observed no significant association between SMA-PM → PCC gamma band connectivity and Berg Balance score in stroke participants. This connectivity-behavioral uncoupling following stroke suggests that although directed communication continues from SMA-PM → PCC, the underlying signal quality is degraded, possibly due to structural changes in the connecting cortico-cortical white matter [24]. Disruptions in functional connectivity after a stroke are primarily determined by the structural disconnection of white matter pathways rather than focal damage to specific cortical gray matter regions [24]. The elevated gamma-band functional connectivity observed in our study likely represents a noisy, low-specificity channel that fails to provide precise information about the upcoming movement for automated balance corrections. The breakdown of automated, narrow-band sensorimotor communication leads to global network reorganization to manage balance control.

### SMA-PM → Contralesional LPFC Connectivity Supports Balance in Stroke

Stroke participants showed stronger SMA-PM → contralesional LPFC connectivity than the reverse direction, whereas healthy adults showed the opposite pattern (a significant Group × Direction interaction). When the automatic sensorimotor network is compromised, as observed in our stroke participants,[90] the balance behavior may depend on reorganized prefrontal-SMA-PM networks [20]. The exchange of information between SMA-PM and prefrontal regions is inherently frequency-specific to implement cognitive control over motor execution [91]. In our study, the stronger connectivity from the SMA-PM → contralesional LPFC was primarily observed when all frequency bands were averaged, suggestive of a loss of standard frequency-specific communication. Importantly, elevated SMA-PM → contralesional LPFC overall connectivity supported better balance during the Berg Balance assessment in stroke participants. This stronger connectivity likely assists with the compensatory cognitive strategy to maintain balance in stroke participants. In stroke participants, the motor/premotor system likely communicates the current balance state and/or the motor plan to contralesional prefrontal regions for performance monitoring and for updating the adaptive balance strategy.

### Ipsilesional LPFC to SMA-PM Connectivity Suggests Increased Executive Influence Over Motor Planning in Stroke

Stroke participants showed stronger Ipsi LPFC→SMA-PM connectivity than in the opposite direction, whereas healthy adults did not show this directional asymmetry (a significant Group × Direction interaction). PFC supports executive control by actively maintaining goal-relevant representations that guide attention, working memory, and action selection, especially when tasks are novel, complex, or require flexible adaptation [92]. PFC maintains goal-relevant representations and can modulate downstream sensory, motor, and association areas via anatomical connections with premotor/SMA pathways [88], so that behavior matches the current goal [92,93]. The LPFC → SMA-PM connectivity likely reflects top-down executive modulation of balance. After a stroke, greater executive influence over motor planning may be required for maintaining an upright stance under challenging balance conditions [90,94]. Similar to contralesional LPFC, the enhanced Ipsilesional LPFC→SMA-PM connectivity was not found to be frequency-specific. Although stroke participants with stronger Ipsi LPFC→SMA-PM connectivity showed better balance numerically, this association did not withstand rigorous statistical correction. This lack of correlation might be due to greater inter-subject variability in ipsilesional LPFC reorganization or a smaller effect size. Stroke participants varied in lesion size, location, and chronicity, potentially introducing heterogeneity that attenuated group-level connectivity-behavior associations.

### Limitations and Future Directions

Several limitations should be considered when interpreting these findings. Our modest sample size means the significant correlation between SMA-PM → contralesional LPFC connectivity and BBS score was driven by a small number of data points. This correlation finding should be confirmed in a larger cohort. Participants with stroke varied in lesion size, location, and time since stroke, and this heterogeneity may have contributed to inter-subject variability in connectivity-behavior associations, mainly for the ipsilesional LPFC → SMA-PM pathway. Furthermore, the correlational nature of connectivity-behavior analyses precludes causal claims. Future studies may include diffusion imaging to corroborate a structural basis for the observed functional pattern. Finally, the connectivity-behavior associations reported here should be understood as linking cortical network activity to global clinical balance ability rather than to task-specific postural performance.

## Conclusion

This study demonstrates that cortical network reorganization after stroke leads to directional connectivity changes that are not uniformly adaptive. Elevated SMA-PM → PCC gamma-band connectivity in stroke was behaviorally uncoupled from balance ability, suggesting that this pathway no longer reliably transmits feedforward information for sensorimotor control of balance. Elevated SMA-PM → contralesional LPFC connectivity was associated with better balance in stroke, suggesting this connection facilitates compensatory balance strategy. The behavioral role of elevated ipsilesional LPFC → SMA-PM connectivity in stroke warrants confirmation in larger samples. These findings suggest that balance rehabilitation after stroke may benefit from approaches that target the quality of sensorimotor communication while reinforcing compensatory contralesional prefrontal pathways.

## Data Availability

Data collected in this study include identifiable clinical information from stroke survivors and are not publicly available due to privacy and ethical restrictions; de-identified data may be available from the corresponding author upon reasonable request and institutional approval.

## ACKNOWLEDGEMENTS

This study was supported by a grant to PJP from the NIH National Center of Neuromodulation for Rehabilitation, the National Institutes of Health Eunice Kennedy Shriver National Institute of Child Health and Human Development (NIH/NICHD) under Grant P2CHD086844 and NIH/NICHD R25HD106896 to PJP and JCV. We want to thank Sheel Shah, Diana Huynh, and Adriel Perez for their assistance with the data collection.

## Notes

### Competing Interest Statement

The authors have declared no competing interest.

### Author Declarations

This study was approved by the Institutional Review Board at the University of Houston.

